# Association of Retinal Age Gap with Arterial Stiffness and Incident Cardiovascular Disease

**DOI:** 10.1101/2022.01.13.22269272

**Authors:** Zhuoting Zhu, Yifan Chen, Wei Wang, Yueye Wang, Wenyi Hu, Xianwen Shang, Huan Liao, Danli Shi, Yu Huang, Jason Ha, Zachary Tan, Katerina Kiburg, Xueli Zhang, Shulin Tang, Honghua Yu, Xiaohong Yang, Mingguang He

**Author notes:** **Corresponding author** Mingguang He, MD PhD, Xiaohong Yang, MD, Honghua Yu, MD PhD. Contributed equally (Zhuoting Zhu, Yifan Chen, Wei Wang).

## Abstract

**Background:** Retinal parameters could reflect systemic vascular changes. With the advances of deep learning technology, we have recently developed an algorithm to predict retinal age based on fundus images, which could be a novel biomarker for ageing and mortality.

**Objective:** To investigate associations of retinal age gap with arterial stiffness index (ASI) and incident cardiovascular disease (CVD).

**Methods:** A deep learning (DL) model was trained based on 19,200 fundus images of 11,052 participants without any past medical history at baseline to predict the retinal age. Retinal age gap (retinal age predicted minus chronological age) was generated for the remaining 35,917 participants. Regression models were used to assess the association between retinal age gap and ASI. Cox proportional hazards regression models and restricted cubic splines were used to explore the association between retinal age gap and incident CVD.

**Results:** We found each one-year increase in retinal age gap was associated with increased ASI (β=0.002, 95% confidence interval [CI]: 0.001-0.003, P<0.001). After a median follow-up of 5.83 years (interquartile range [IQR]: 5.73-5.97), 675 (2.00%) developed CVD. In the fully adjusted model, each one-year increase in retinal age gap was associated with a 3% increase in the risk of incident CVD (hazard ratio [HR]=1.03, 95% CI: 1.01-1.06, P=0.012). In the restricted cubic splines analysis, the risk of incident CVD increased significantly when retinal age gap reached 1.21 (HR=1.05; 95% CI, 1.00-1.10; P-overall <0.0001; P-nonlinear=0.0681).

**Conclusion:** We found that retinal age gap was significantly associated with ASI and incident CVD events, supporting the potential of this novel biomarker in identifying individuals at high risk of future CVD events.

## Introduction

Amongst non-communicable diseases, cardiovascular disease (CVD) is the leading cause of morbidity and mortality globally. From 2005 to 2015, global CVD deaths increased by 12.5% to 17.9 million per year.^1^ Approximately one in three deaths worldwide can be attributed to CVD.^2^ Driven by an ageing and growing population, the burden of CVD is projected to increase further.

Prevention and early detection of CVD are crucial in reducing CVD morbidity and mortality.^3^ Many CVD risk calculators have thus been developed, including the Cardiovascular Risk Score (QRISK), Systemic Coronary Risk Evaluation (SCORE), and Framingham risk score.^4-7^ Nevertheless, these risk prediction models are limited by lack of precision, information bias for questionnaire-based models, and the involvement of invasive procedures.^6, 8-10^

The retina is highly vascular and easily accessible to non-invasive imaging and assessments and can serve as a surrogate measure of the health of the systemic vasculature. Collective evidence has demonstrated that fundus images contain valuable information on cardiovascular health. Changes in retinal vascular parameters derived from fundus images, such as wider retinal venules and narrower retinal arterioles, have been found to be independent predictors of future CVD risk.^11-18^ The advent of deep learning (DL) has greatly improved the efficacy and accuracy of extracting salient features from images. Recent studies have demonstrated the predictive value of retinal fundus images on CVD risk profiles and atherosclerosis via DL algorithms (DLA).^19-21^

To maximally utilize available information from the retina, we have developed a DLA that can accurately predict an individual’s chronological age from fundus images. The retinal age gap (defined as fundus image predicted age minus chronological age) was found in our previous study to be associated with an increased risk of mortality, further lending credence to its potential as a robust biomarker of ageing.^22^ Age is one of the most important risk factors for atherosclerosis and CVD, with CVD risks increasing exponentially with age.^23, 24^ Therefore, we hypothesized that retinal age gap may have the potential to be a novel biomarker for future CVD risk.

Herein, we aimed to investigate the association of the retinal age gap with the arterial stiffness index (ASI). As the arterial system ages, large arteries undergo progressive elastic and collagenous remodeling that result in increased stiffness. ASI is an accepted, non-invasive measure of cardiovascular health and an indicator of subclinical CVD.^25^ Further, we also investigated the relationship of the retinal age gap with the future risk of incident CVD based on the UK Biobank study.

## Methods

### Study population

The UK Biobank is a large-scale prospective study with over 500,000 participants aged 40-69 years recruited between 2006 and 2010. This study has collected extensive phenotypic and genotypic data of each participant with their informed consent. All participants completed questionnaires on their lifestyle, environment and medical history, underwent physical and functional measurements, and provided blood, urine and saliva samples. Ophthalmic examinations including retinal photographs were introduced in 2010. Health-related events were recorded via linkage with Hospital Episode Statistics (HES) and death registers. A detailed description of the UK Biobank data and protocols can be found elsewhere.^26^

### Standard Protocol Approvals, Registrations, and Patient Consents

Access to the UK Biobank data was granted after registration. The application ID was 62443. The North West Multi-centre Ethics Committee granted ethical approval to the UK Biobank (11/NW/0382). The current study operates in accordance with the principles of the Declaration of Helsinki, with written informed consent from all participants, under the UK Biobank application number 62489.

### Fundus photography

The physical measures of ophthalmic examination included LogMAR visual acuity, autorefraction and keratometry (Tomey RC5000, Tomey GmbH, Nuremberg, Germany), intraocular pressure (IOP, Ocular Response Analyzer, Reichert, New York, USA), and paired retinal fundus and optical coherence tomography imaging (OCT, Topcon 3D OCT 1000 Mk2, Topcon Corp, Tokyo, Japan). A 45-degree non-mydriatic and non-stereo fundus image centered to include both the optic disc and the macula was taken for each eye. In total, 131,238 images from 66,500 participants collected at the baseline were obtained from the UK Biobank study.

### Arterial stiffness

The ASI was derived from the analysis of the digital volume pulse, an indirect method to assess AS peripherally. ASI data were collected at baseline via the PulseTrace PCA 2 (CareFusion), which uses finger photoplethysmography to assess the pulse waveform using an infrared sensor placed on the index finger of the participant’s dominant hand. The pulse waveform comprises a systolic peak and a second diastolic peak, and the transit time (peak-to-peak time [PPT]) between the 2 peaks is related to the time it takes for the pulse wave to travel through the peripheral arterial tree. The length of this arterial tree path is proportional to a person’s height (h), enabling the calculation of an index of large artery stiffness using the formula ASI[=[h[/[PPT. The ASI was natural log transformed.

### Cardiovascular disease ascertainment

To ascertain CVD events, the date of the first known myocardial infarction or stroke was identified via linkage with hospital admission data in England, Scotland and Wales, the national death register data and self-reporting. Myocardial infarction was defined by codes 410, 411, 412.X, 429.79 in the 10^th^ edition of International Classification of Diseases (ICD-9), and codes I21, I22, I23, I24.1 or I25.2 in the 10th edition of ICD-10. Stroke was defined by ICD-9 codes 430.X, 431.X, 434.X, 434.0, 434.1, 434.9, 436.X, and ICD-10 codes I60, I61, I63, or I64.X. History of CVD events was defined as the occurrence of CVD events prior to the baseline examination. Incident CVD events was defined as the first occurrence of CVD events during the follow-up period, from the baseline examination to 29 Feb 2016, or the first occurrence of either myocardial infarction or stroke. Participants with a history of CVD events were excluded from the analysis of the association between retinal age gap and incident CVD.

### Deep learning model for age prediction

A total of 80,169 images from 46,969 participants passed the image quality check. The details of quality check have been documented elsewhere.^27^ Briefly, the fundus images were rated by two ophthalmologists as good, usable or reject based on a three-level quality rating system, which took into account four quality indicators including blurring, uneven illumination, low-contrast, and artifacts. Among 46,969 participants, 11,052 participants did not report any previous disease. A total of 19,200 fundus images from 11,052 participants without any past medical histories at baseline were used to build the DL model for age prediction. Images from both eyes if available were used to maximize the volume of data available. Among the remaining 35,917 participants, 35,541 with available ASI data and its value within 4 standard deviations (SDs) of the mean were included to investigate the association between retinal age gap and ASI. A total of 33,817 participants who had no history of CVD events at baseline were included to investigate the effects of retinal age gap on the risk of incident CVD events. Images from the right eye were used to calculate the retinal age and were replaced by images from the left eye if not available.

The development and validation of the DL model for age prediction were described in details elsewhere.^22^ Briefly, all fundus images were preprocessed and fed into a DL model using a Xception architecture. Data augmentation was performed during training using random horizontal or vertical flips. The algorithm was optimized using stochastic gradient descent. To avoid overfitting, we implemented a dropout of 0.5 and carried out early stopping when validation performance did not improve for 10 epochs. The DL model was trained and validated using five-fold cross-validation. The performance of the DL model was calculated, including the mean absolute error (MAE) and the correlation between predicted retinal age and chronological age. We then retrieved attention maps from the DL model using guided Grad-CAM, which highlights pixels in the input image based on their contributions to the final evaluation.

The trained DL model was able to achieve a strong correlation of 0.80 (P<0.001) between predicted retinal age and chronological age, with an overall MAE of 3.55 years. The retinal age gap was defined as the difference between the retinal age predicted by the DL model and the chronological age. The retinal age gap would be positive for an ‘older’ appearing retina compared to chronological age, while a ‘younger’ appearing retina would have a negative retinal age gap.

### Covariates

Confounding factors included baseline age, sex, ethnicity (recorded as white or non-white), Townsend deprivation indices (an area-based proxy measure for socioeconomic status), education attainment (recorded as college/university degree and above, or others), smoking status (recorded as current/previous or never), drinking status (recorded as current/previous or never), physical activity level (recorded as above physical education recommendation or not), history of CVD events (for the association between ASI and the retinal age gap), metabolic syndrome, and general health status (recorded as excellent/good and fair/poor). Metabolic syndrome was defined as the presence of three or more of the following factors: unhealthy waist circumference, hypertension, dyslipidemia, hypertriglyceridemia, and hyperglycemia. We used established reference values for women (≥88 cm) and men (≥102 cm) as cutoff values to define unhealthy waist circumference. Hypertension was defined as a systolic blood pressure of 130 mm Hg or above or a diastolic blood pressure of 85 mm Hg or above or taking antihypertensive drugs. Dyslipidemia was defined as a high-density lipoprotein cholesterol level of less than 40 mg/dL among men and less than 50 mg/dL among women. Hypertriglyceridemia was defined as triglyceride levels of 150 mg/dL or greater. Hyperglycemia was defined as fasting blood glucose levels greater than 110 mg/dL or taking medications/insulin for diabetes.

### Statistical analyses

Descriptive statistics, including means and standard deviations (SDs), numbers and percentages, were used to report baseline characteristics of study participants. Linear regression models considering retinal age gap as a continuous linear term were fitted to investigate the association between a one-year increase in retinal age gap and ASI. We then investigated the associations between retinal age gaps of different quintiles and ASI. For the logistic regression models, we dichotomized the outcomes into ASI equal to/above or below the median ASI. An ASI equal to or above the median ASI was defined as a severe ASI. We investigated the effect of a one-year increase in retinal age gap on the odds ratio (OR) of having a severe ASI. The following covariates – baseline age, sex, and ethnicity (model I); additional Townsend deprivation indices, educational level, smoking status, drinking status, physical activity level, history of CVD events, metabolic syndrome, and general health status (model II) were adjusted for in the regression models.

Cox proportional hazards regression models were used to explore the relationship between retinal age gap (continuous or quintiles) and incident CVD. We adjusted Cox proportional hazards regression models for the following covariates – baseline age, sex, and ethnicity (model I); additional Townsend deprivation indices, educational level, smoking status, drinking status, physical activity level, history of CVD events, metabolic syndrome, and general health status (model II); additional ASI (III).

The proportional hazards assumption for each variable included in the Cox proportional hazards regression models were assessed graphically. Restricted cubic spline analyses were performed to further assess the association between retinal age gap and incident CVD. Five knots were placed at equal intervals across the distribution of the retinal age gap. Retinal age gap of zero years was used as the reference. All variables were found to meet the assumption. Variance inflation factors (VIF) procedure was used to test collinearities for all variables and all covariables’ VIF were less than 2 (mean: 1.16). A two-sided p-value of <0.05 indicated statistical significance. Analyses were performed using Stata (version 13, StataCorp, Texas, USA).

## Results

### Study sample

The baseline characteristics of 35,541 included participants were shown in Table 1. The mean age was 56.8 ± 8.04 years, and 55.6% were females.

**Table 1.**
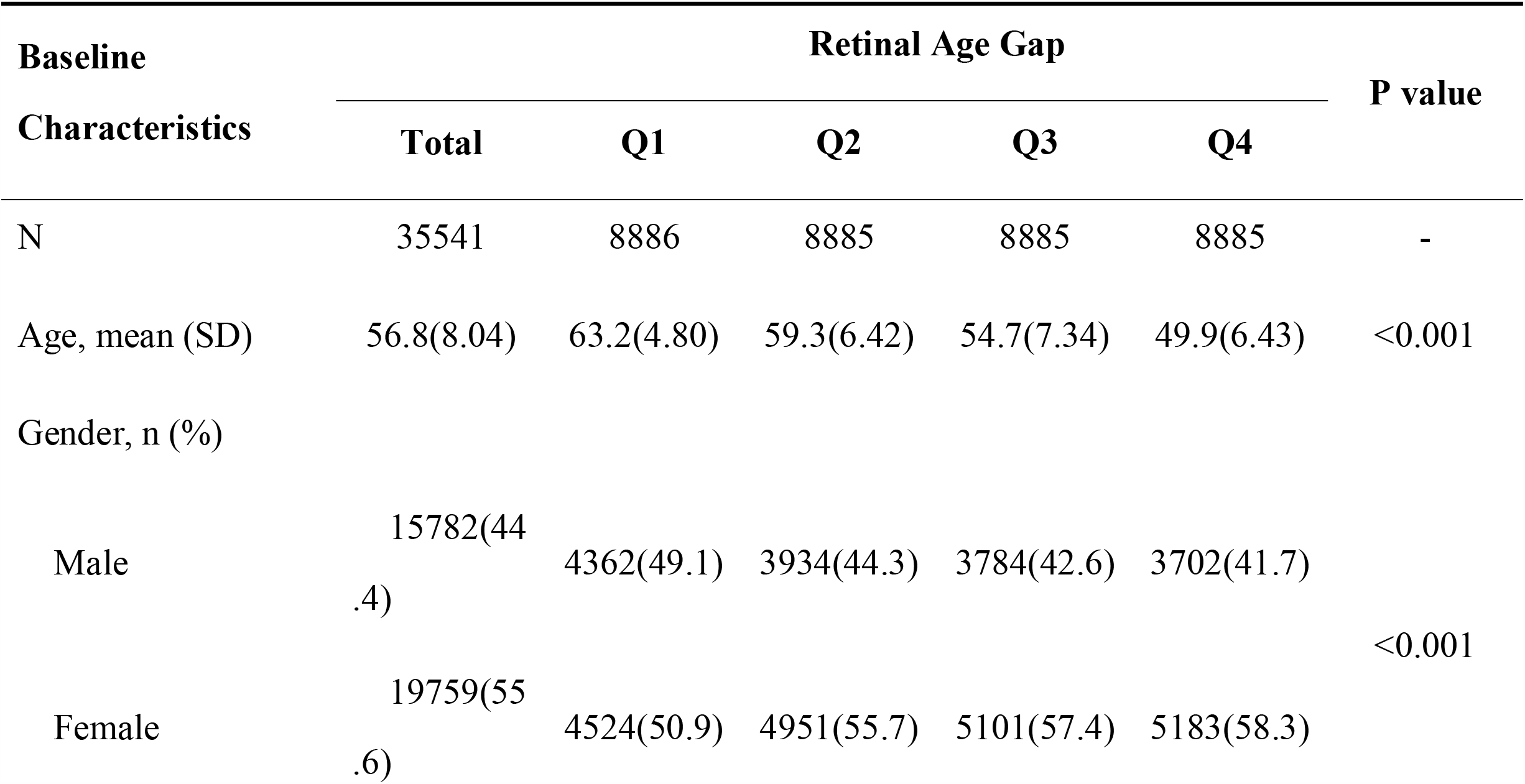
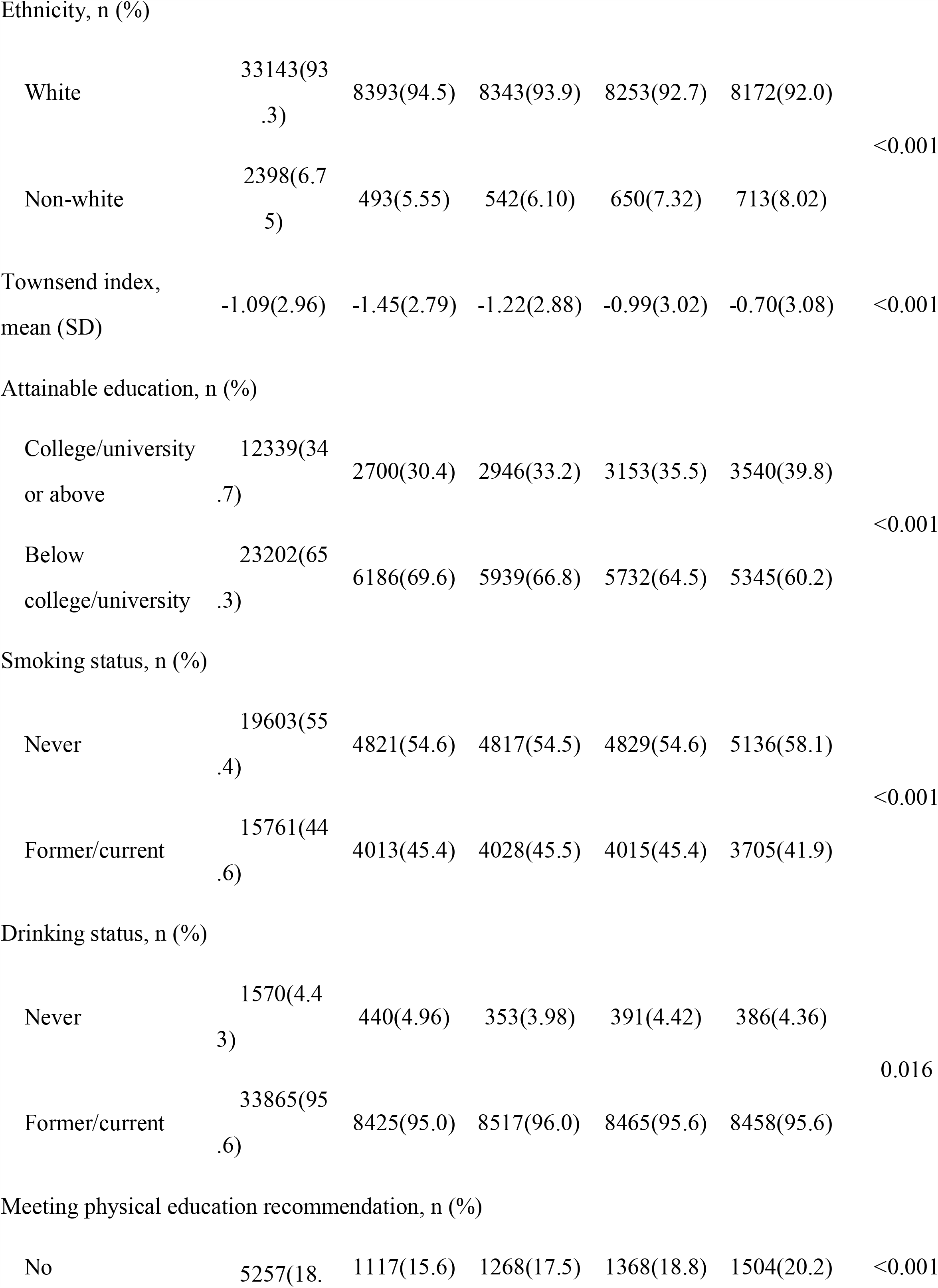
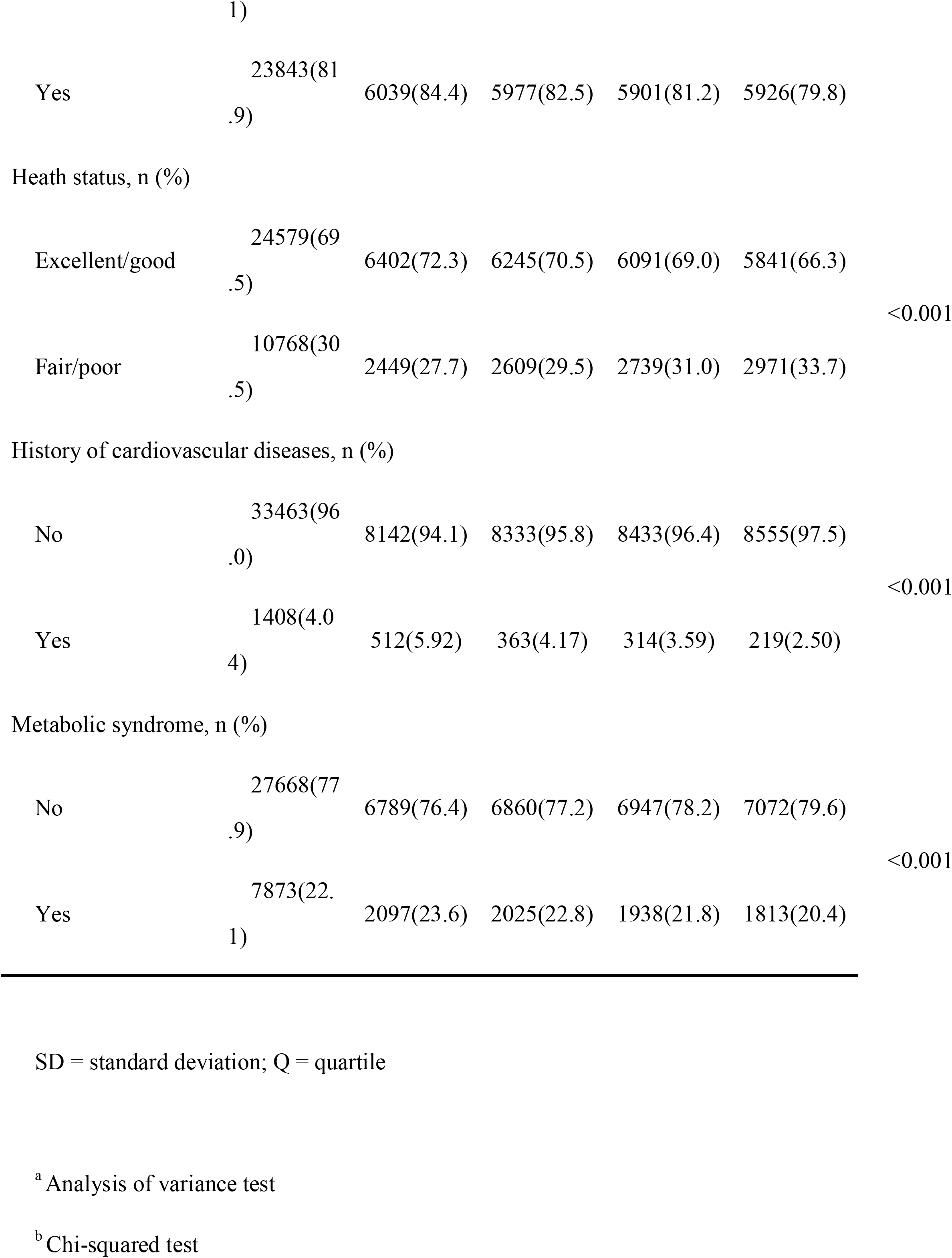
Baseline Characteristics of Study Participants Stratified by Quartiles of Retinal Age Gap

### Retinal age gap and arterial stiffness index

Linear regression models were implemented for the investigation of the retinal age gap and continuous ASI. We found that a one-year increase in the retinal age gap was associated with a significant increase in ASI with a β coefficient of 0.002 (95% confidence interval [CI]: 0.001-0.003, P<0.001) after adjusting for all included confounders. When the participants were divided into quartiles by their retinal age gaps, those in the second (β=0.013, 95% CI: 0.002-0.023, P=0.018), third (β=0.018, 95% CI: 0.007-0.029, P=0.002) and fourth (β=0.017, 95% CI: 0.004-0.030, P=0.009) quartiles had significantly higher ASI values compared to those in the first quartile (Table 2).

**Table 2.**
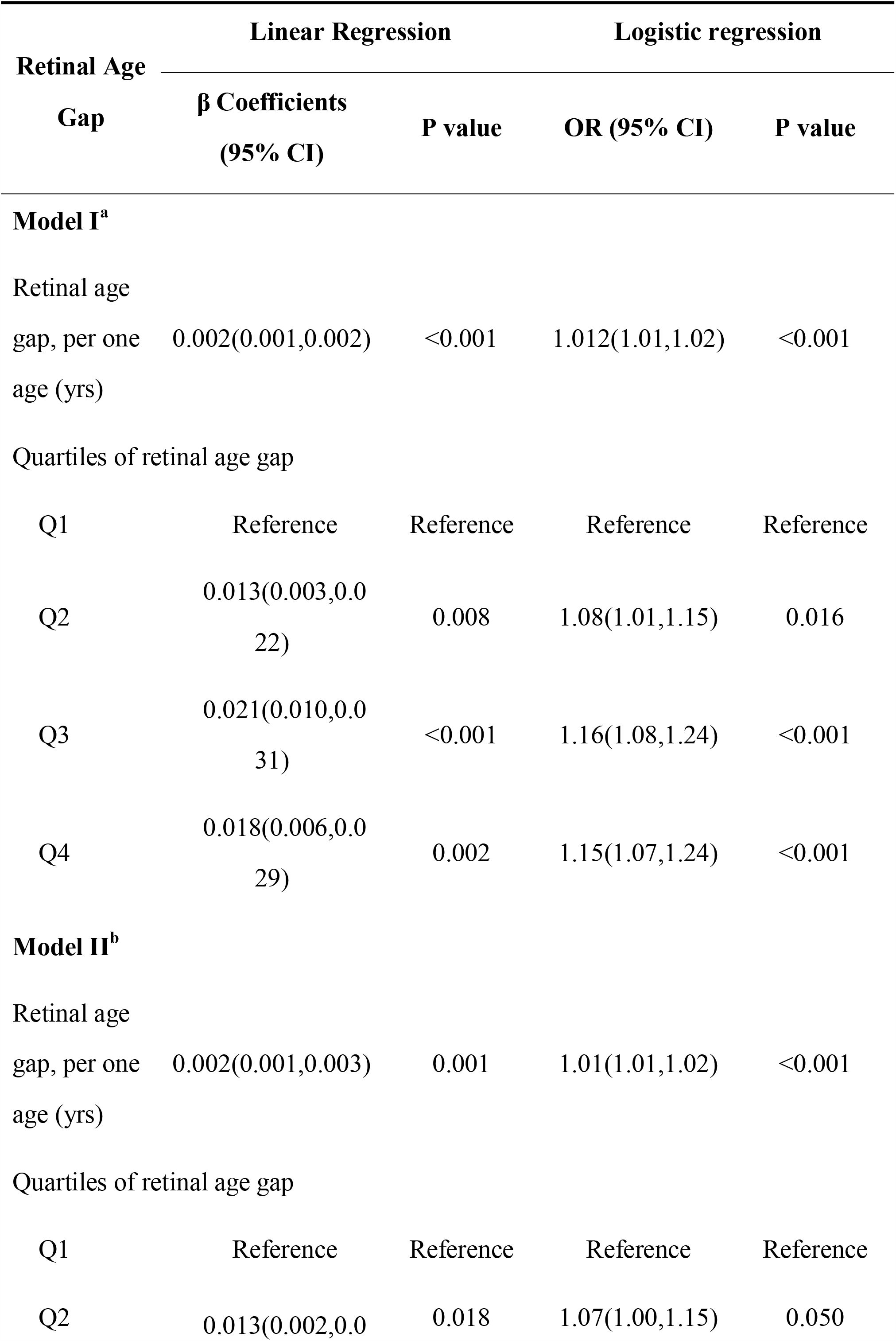
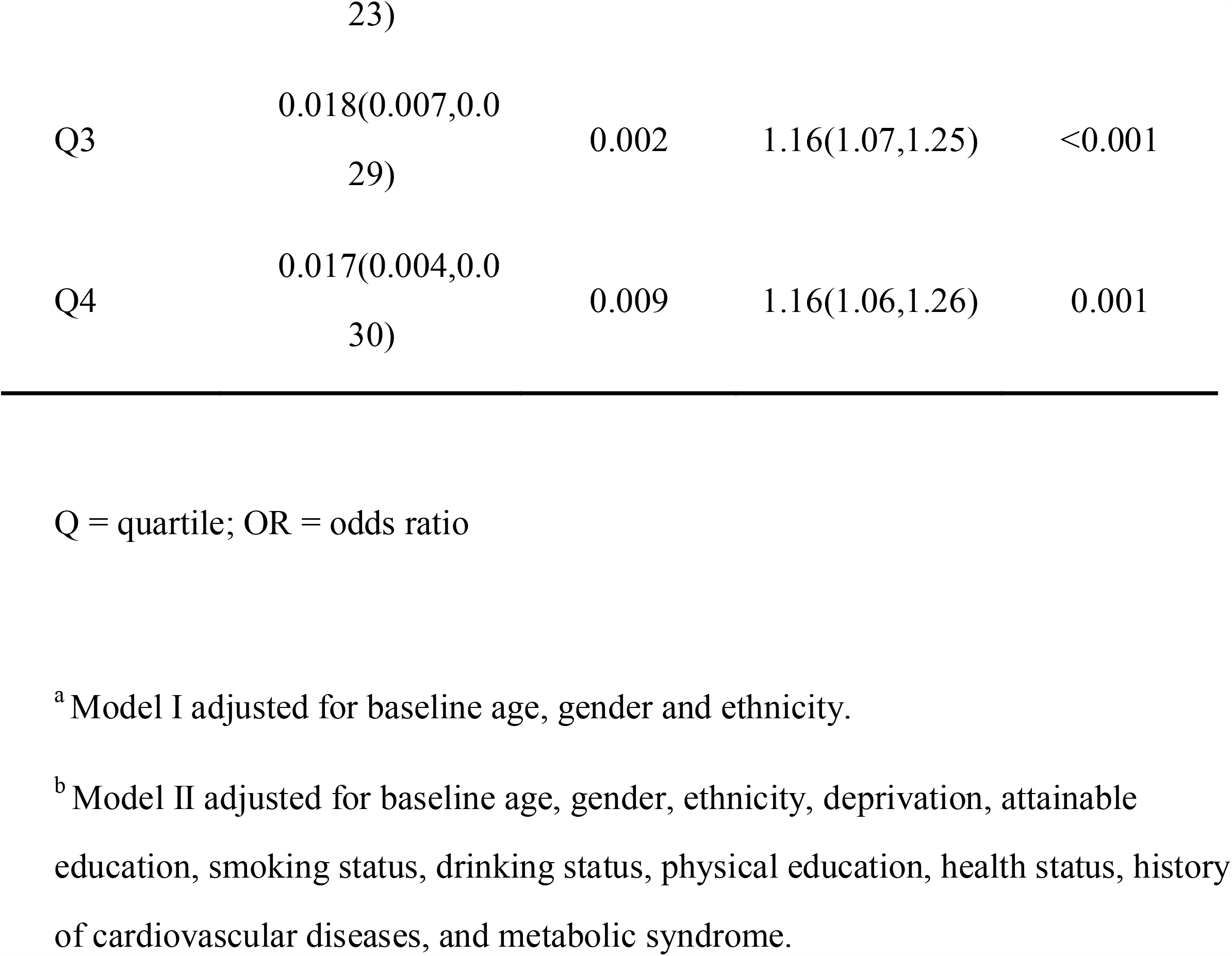
β Coefficients and Odds Ratio (OR) for the Association between Retinal Age Gap and Arterial Stiffness Index

Considering ASI as a categorical variable, logistic regression models indicated higher odds of having severe ASI with a larger retinal age gap. The OR of having severe ASI was 1.01 (95% CI: 1.01-1.02) with each one-year increase in the retinal age gap when adjusting for all included confounders. It was shown that participants in the third (OR=1.16, 95% CI: 1.07-1.25, P<0.001) and fourth (OR=1.16, 95% CI: 1.07-1.26, P=0.001) quartiles had significantly higher odds of having severe ASI compared to those in the first quartile.

### Retinal age gap and incident CVD

A total of 33,817 participants had no history of CVD events at baseline. The median duration of follow-up was 5.83 years (interquartile range [IQR]: 5.73-5.97). During the follow-up period, a total of 675 (2.00%) participants had incident CVD events. After adjusting for confounders, each one-year increase in retinal age gap predicted a 3% increase in the risk of incident CVD (hazard ratio [HR]=1.03, 95% CI: 1.01-1.06, P=0.012). This independent predictive value remained significant when ASI was incorporated into the fully adjusted model (HR=1.03, 95% CI: 1.01-1.06, P=0.009). Compared to participants with retinal age gaps in the first quartile, those with retinal age gaps in the second (HR=1.08, 95% CI: 0.86-1.36, P=0.481) and third quartiles (HR=1.10, 95%CI: 0.84-1.43, P=0.495) had similar risks of developing incident CVD, while the risk was markedly increased in the fourth quantile (HR=1.54, 95%CI: 1.12-2.11, P=0.007) (Table 3). In the restricted cubic splines analysis, there were four knots in total. When retinal age gap=0 was used as reference, the risk of incident CVD increased significantly when the retinal age gap reached 1.21 (HR=1.05; 95% CI, 1.00-1.10; P-overall <0.0001; P-nonlinear=0.0681, Figure 1).

**Table 3.**
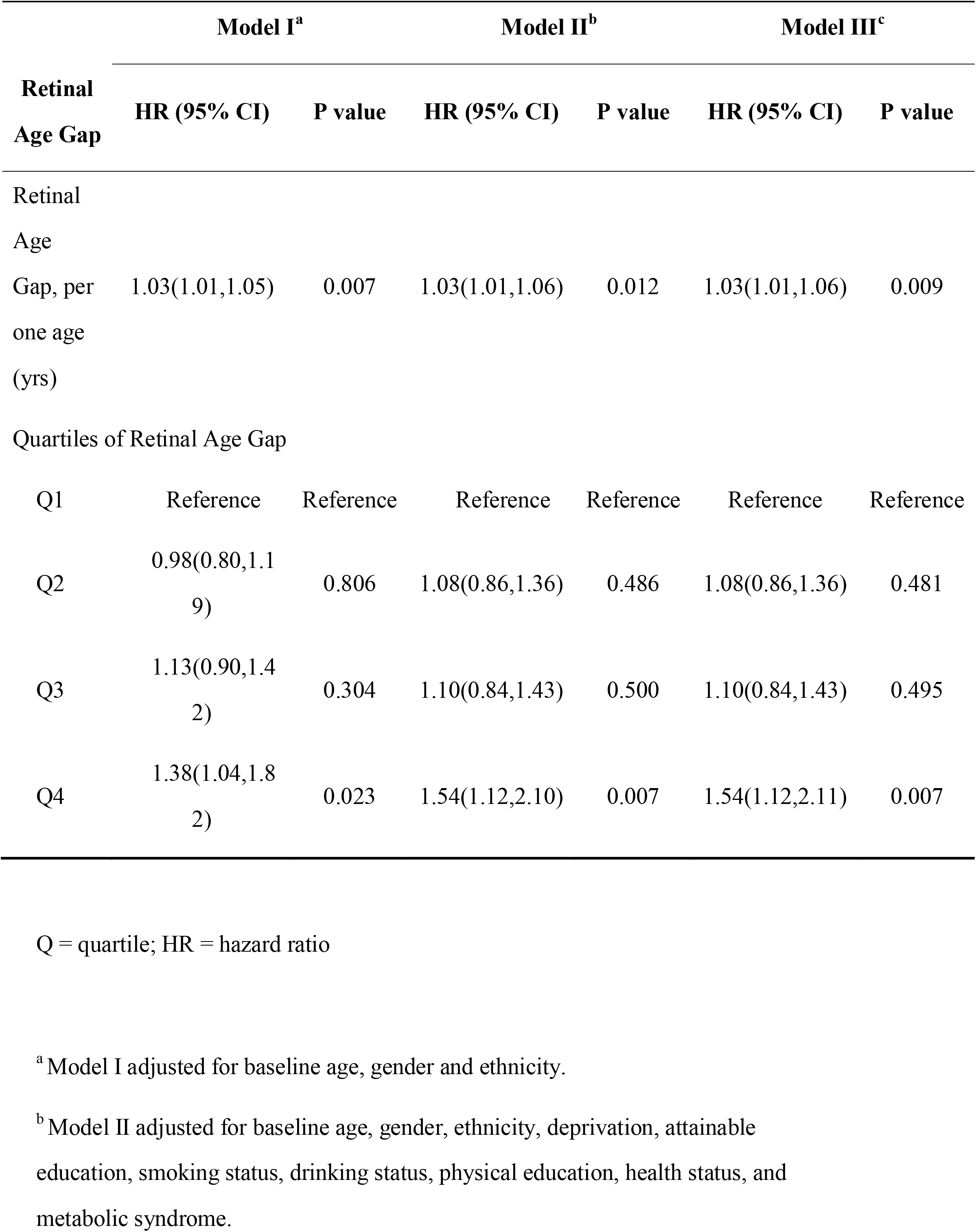
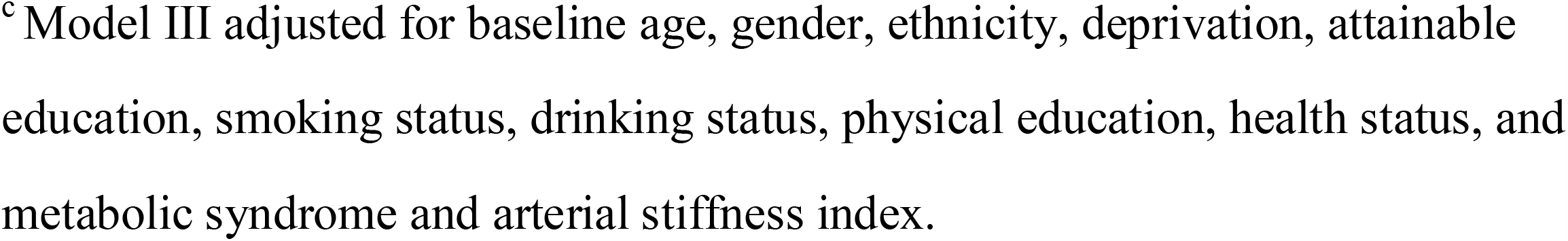
The Association between Retinal Age Gap and Incident Cardiovascular Diseases

**Figure 1:**
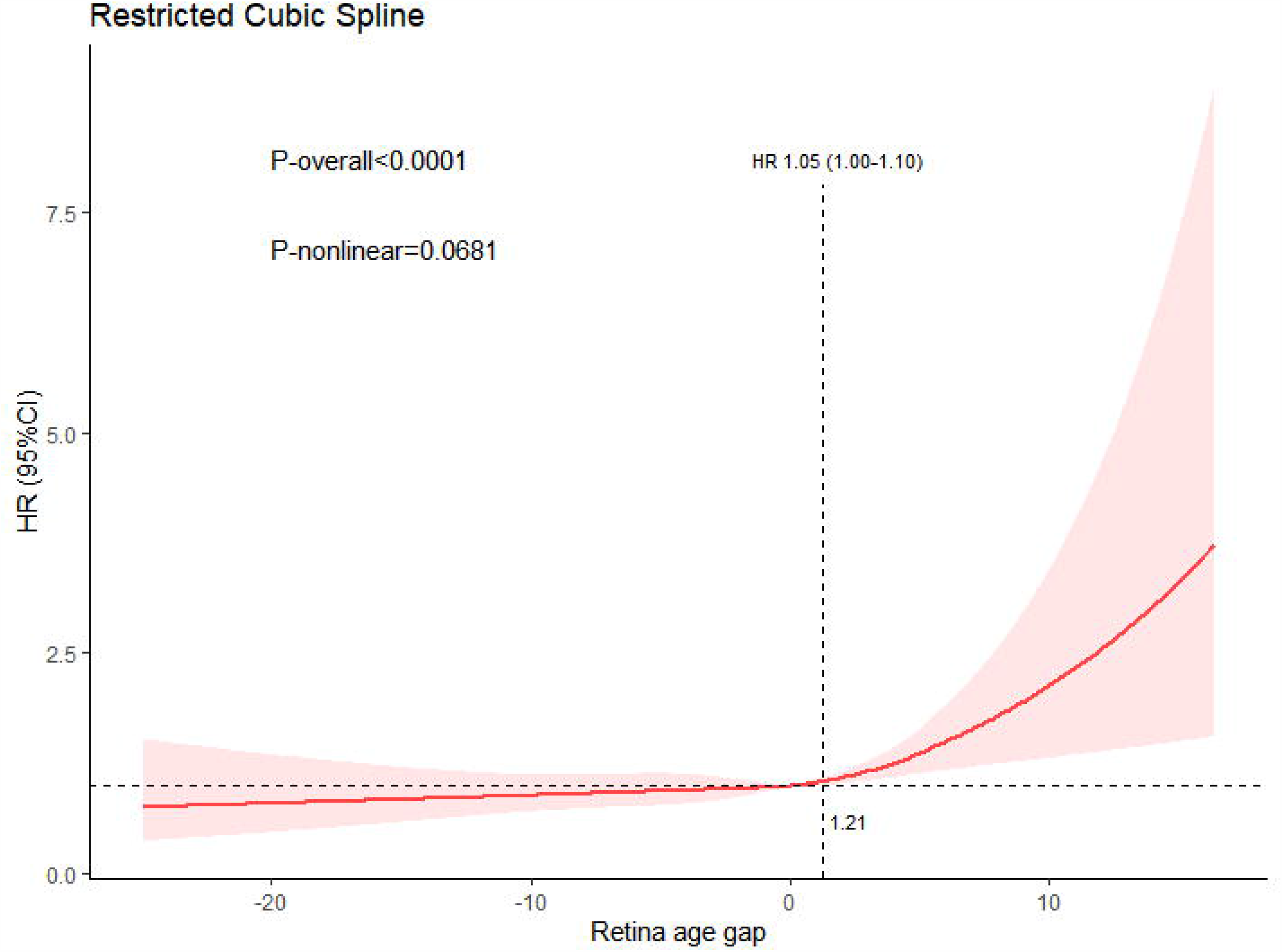
Non-linear association between retinal age gap and incident CVD. Restricted cubic spline analyses were performed to assess the association between retinal age gap and incident CVD allowing non-linearity. Five knots were placed at equal intervals across the distribution of the retinal age gap. Retinal age gap of zero years was used as the reference. Overall association between retinal age gap was observed (P_overall_ < 0.001). A J-shaped curve was observed for the association between retinal age gap and incident CVD, where the association was significant only when retinal age gap exceeded 1.21 years.

## Discussion

In the present study, we found that retinal age gap was significantly associated with ASI. In addition, the retinal age gap was found to be a predictor of future risk of incident CVD, independent of traditional risk factors, as well as ASI.

Our finding of the association between retinal age gap and ASI was consistent with previous studies, where strong associations between quantitatively assessed retinal abnormalities (e.g., retinal arteriolar narrowing and venular widening) and ASI were demonstrated.^28-31^ As a clinical parameter, ASI is used to estimate the stiffness of blood vessels and an early indicator of atherosclerosis.^32^ Higher ASI may indicate microcirculation dysfunction and target organ damage.^33, 34^ Moreover, previous research has demonstrated ASI is a strong predictor of future CVD events.^35^ The consistent evidence suggested that the retinal age gap might be a novel biomarker of subclinical CVD.

In addition, retinal age gap was found to be an independent biomarker for future incident CVD events in this study. This association remained significant even after adjusting for baseline ASI. In keeping with our findings, a large number of previous studies have identified valuable information from fundus images including retinal microvascular signs (e.g., narrower retinal arteriolar caliber and wider retinal venular caliber) and geometric parameters (e.g., vessel tortuosity and bifurcation angle), as predictors for future CVD events and mortality.^12, 14, 36-46^ Recently, with the advances in DL technologies, characteristics derived from retinal fundus images could be automatically extracted and learned with high efficiency for the prediction of CVD profiles and events.^19-21^ Of note, the attention maps of our DL model^22^ mainly highlighted the areas around the retinal vessels, suggesting that the vasculature and their surrounding areas were the key areas for identification of age. This spatial validation of the salience of the retinal vasculature in fundus photographs further enhances its role as a biomarker for the health of the systemic vasculature.

The pathophysiological mechanisms underlying our finding that CVD profiles and future events could be predicted from retinal images warrant further investigation. The similarities and interplay between the vasculature of the eye and the heart are likely to be the fundamental contributors.^47^ As one of the predominant risk factors, ageing might result in similar pathological changes in the vasculature of these two organs. Firstly, excessive oxidative stress, lipid accumulation, and chronic inflammation caused by ageing can consequently lead to endothelial dysfunction, which is a crucial process implicated in both the dysregulation of retinal blood flow and most CVD.^48, 49^ Secondly, various age-related comorbidities such as hypertension, diabetes, and atherosclerosis, were found to be risk factors of CVD while simultaneously associated with structural vascular changes in the retina.^47, 50^

The current study found that retinal age gap may be a promising novel biomarker for future CVD risks with several important clinical implications. Screening for eye diseases through fundus photography is efficient, non-invasive and cost-effective, and recommended for diabetic patients as part of annual screening programmes.^51^ The evaluation of retinal age gap by a DLA can be easily integrated into ocular screening programmes for the early detection and monitoring of CVD with enhanced cost-effectiveness. Moreover, with the rapid development of portable electronic devices, retinal age gap may in the future be derived from smartphone-based retinal cameras and used as a point of care assessment of CVD risk; naturally, the use of non-standardised image acquisition systems will require validation for its accuracy in future studies. Clinical application of retinal age gap may achieve an instant evaluation of systemic vasculature conditions and improve accessibility to CVD risk evaluation. It may also facilitate the early referral of patients with high CVD risk for prompt cardiology assessment and subsequent preventative interventions.

The strengths of the current study include a large sample size, relatively long duration of follow-up, the standardized protocol used for capturing fundus images, and adjustment for a wide range of confounding variables. Some limitations also remain. Firstly, the retinal age gap was calculated based on fundus images captured at a specific time point. Dynamic changes in the ageing of the retina over time may be a better predictor for the future risk of CVD. Secondly, we could not exclude the possibility of residual confounding effects.

## Conclusion

We found that retinal age gap generated by a DLA was positively associated with ASI and risk of future incident CVD. This study suggested that retinal age gap can be utilized as a novel biomarker for CVD risk evaluation, early detection and monitoring. Future studies are needed to validate our findings in different populations.

## Data Availability

All data produced in the present study are available upon reasonable request to the authors

https://www.ukbiobank.ac.uk/enable-your-research/apply-for-access

## Contributors

Study concept and design: Zhu ZT, Chen YF, Wang W, He MG, Yang XH.

Acquisition, analysis, or interpretation: All authors.

Drafting of the manuscript: Zhu ZT, Chen YF.

Critical revision of the manuscript for important intellectual content: Wang W, Hu WY, Kiburg K, Zhu ZT, He MG, Yang XH.

Statistical analysis: Zhu ZT.

Obtained funding: He MG, Yang XH.

Administrative, technical, or material support: Zhu ZT, Wang W, He MG, Yang XH.

Study supervision: He MG, Yang XH, Yu HH.

## Acknowledgments

This present work was supported by the NHMRC Investigator Grant (APP1175405), Fundamental Research Funds of the State Key Laboratory of Ophthalmology, National Natural Science Foundation of China (82000901, 82101173), Project of Investigation on Health Status of Employees in Financial Industry in Guangzhou, China (Z012014075), Science and Technology Program of Guangzhou, China (202002020049), Research Foundation of Medical Science and Technology of Guangdong Province (B2021237). Professor Mingguang He receives support from the University of Melbourne through its Research Accelerator Program and the CERA Foundation. The Centre for Eye Research Australia (CERA) receives Operational Infrastructure Support from the Victorian State Government.

